# Psychopathology in mothers of children with pathogenic Copy Number Variants

**DOI:** 10.1101/2020.09.24.20201053

**Authors:** Maria Niarchou, Adam C. Cunningham, Samuel J.R.A. Chawner, Hayley Moulding, Matthew Sopp, IMAGINE-ID, Jeremy Hall, Michael J. Owen, Marianne van den Bree

## Abstract

Caring for children with pathogenic neurodevelopmental Copy Number Variants (CNVs) (i.e., deletions and duplications of genetic material) can place a considerable burden on parents, and their quality of life. Our study is the first to examine the frequency of psychiatric diagnoses in mothers of children with CNVs compared to the frequency of psychiatric problems in age-matched mothers from a large community study. 268 mothers of children with CNVs had higher frequency of depression compared to the 2,680 age-matched mothers (p<0.001). Mothers of children with CNVs reported higher frequency of anorexia, bulimia, alcohol abuse and drug addiction compared to the age-matched mothers from the community sample. The frequency of depression arising after the birth of the index child was similar between the two groups (48% in mothers of children with CNVs vs. 44% in mothers of the community sample, p=0.43), but mothers of children with CNVs had higher frequency of anxiety (55%) compared to mothers from the community sample (30%, p=0.03). Our study highlights the need for health-care providers to devise treatment plans that not only focus on meeting the child’s needs, but also assessing and if needed, addressing the mental health needs of the parent.

## Introduction

Recent genetic advances have led to the detection of deletions or duplications of chromosomal regions, otherwise known as Copy Number Variants (CNVs). Certain relatively rare Copy Number Variants (CNVs) have been shown to increase risk for a number of neurodevelopmental and psychiatric conditions, including Attention Deficit Hyperactivity Disorder, and Autism Spectrum disorder (Chawner et al., 2019; Niarchou et al., 2014) and other health problems (Ionita-Laza, Rogers, Lange, Raby, & Lee, 2009).

The multitude of physical and health comorbidities associated with pathogenic CNVs in children can place a considerable burden on parental caregiving (Baker, Devine, Ng-Cordell, Raymond, & Hughes, 2020). For example, there is evidence that caregiving a child with a developmental disorder can be taxing on the parent in terms of their emotional (e.g., (Y. Mori, Downs, Wong, & Leonard, 2019)) and physical well-being (e.g., (Laurvick et al., 2006)). Research in mothers has identified that mothers with a child with a developmental disability were more likely to experience difficulties coping (Minnes, Perry, & Weiss, 2015), poor emotional well-being relative to general female populations (Yuka Mori, Downs, Wong, Heyworth, & Leonard, 2018), and more child behavioural problems (e.g.,(Totsika, Hastings, Emerson, Lancaster, & Berridge, 2011)).

Taking into account the ‘diagnostic odyssey’ that sometimes parents of children with CNVs have to go through (Anderson, Elliott, & Zurynski, 2013), that in turn can impact the parents’ quality of life (Lingen et al., 2016), as well as the burden of caregiving on the parents placed by the multitude of physical and health comorbidities associated with pathogenic CNVs, there is paucity of research to examine the frequency of psychiatric problems in the parents of children with CNVs. Previous research has indicated high emotional distress in parents of children with CNV (Baker et al., 2020). However, our study is the first to examine the frequency of psychiatric diagnoses in mothers of children with CNVs in comparison with the frequency of psychiatric problems in age-matched mothers from a large community study. We hypothesized that mothers of children with CNVs have more psychiatric problems compared to mothers from the general population.

## Methods

### Samples

#### Mothers of children with CNV

Recruitment of the children with a CNV was through 14 genetics clinics across the UK and the charities Max Appeal, Unique and the 22Crew, as well as social media and word of mouth.

Children’s CNV genotypes were established from medical records as well as in-house genotyping at the Cardiff University MRC Centre for Neuropsychiatric Genetics and Genomics using microarray analysis. Considering our overarching goal of examining the psychopathology of mothers of children with CNVs, we considered it appropriate to also include all children who took part in face-to-face assessments in these studies, meaning that a variety of CNVs was represented.

272 mothers with children with a CNV (mean age 41.2 years (SD=9.8) at the time of assessment and mean age 30.3 years (SD=7.0) when their child was born were recruited from the ExperienCes of people witH cOpy number variants study (ECHO) (see: https://www.cardiff.ac.uk/mrc-centre-neuropsychiatric-genetics-genomics/research/themes/developmental-psychiatry/echo-study-cnv-research), and the IMAGINE-ID study (http://www.imagine-id.org/).

For 53% of our sample (N=145) we also had information from the parents on whether the CNV was de novo or was inherited. 26% of the mothers (N=38/145) were CNV carriers.

Our study was approved by the National Health Service Wales Research and NHS London Queen Square research ethics committees. Parents provided informed written consent for their participation.

#### Mothers of children from a community sample

2,680 mothers, matched for their age and the age of their child to the mothers of children with CNV sample, were recruited from the Avon Longitudinal Study of Parents and Children (ALSPAC). ALSPAC (http://www.bristol.ac.uk/alspac/) is a population-based cohort that was established in the southwest of England. Pregnant women resident in Avon, UK with expected dates of delivery 1st April 1991 to 31st December 1992 were invited to take part in the study. The initial number of pregnancies enrolled is 14,541 (for these at least one questionnaire has been returned or a “Children in Focus” clinic had been attended by 19/07/99). Of these initial pregnancies, there was a total of 14,676 foetuses, resulting in 14,062 live births and 13,988 children who were alive at 1 year of age (Boyd et al., 2012; Fraser et al., 2012).

Ethical approval was provided by the Local Research Ethics Committees and the ALSPAC’s Law and Ethics Committee. Parents provided informed written consent and were allowed to withdraw at any time.

### Measures

#### Mothers of children with CNV

Psychopathology in the mothers of children with CNV was assessed using the Parental Psychopathology section of the parent version of the Child and Adolescent Psychiatric Assessment (CAPA) (Angold et al., 1995). The CAPA is a semi-structured interview that generates categorical diagnoses as well as symptom counts of childhood psychopathology. The parental psychopathology section includes questions about parent’s own psychopathology. Stem questions enquire about whether the parent has ever had depression, anxiety, panic, eating disorder, Obsessive Compulsive Disorder (OCD), or psychosis, or problems with alcohol or drugs. If the parent answered yes to any of these questions, subsequent questions were asked enquiring about the severity of the problem (i.e., whether the subject sought treatment, received medication, or was hospitalized).

#### Mothers from community sample

Various questionnaires were administered to mothers at different time points of their child’s development. To enable comparisons with the ECHO study, we selected questions included in the questionnaires that 1) were phrased either identically or as close as possible to the questions asked in the ECHO and IMAGINE-ID studies and 2) were administered when the children and the mothers in the ALSPAC study were within a similar age range to the children and the mothers of the ECHO and IMAGINE-ID studies, in order to facilitate matching (see Data Analysis below). For these reasons, we selected the mental health related questions that were included in the ‘Lifestyle and health of mother’ questionnaire (Section A – standard ALSPAC health related questions) that was administered to mothers when their child was 8 years old, as the age-range of the mothers and the children was similar to the age-range of the mothers and children in the ECHO study. Taking into account that in the ECHO and IMAGINE-ID studies the questions about depression included treatment and hospitalization, we compared this question with the question ‘Have you ever had severe depression’ which was included in the above questionnaire that was administered to mothers when their child was 11 years old.

#### Data analysis

We ran descriptive statistics using R version 3.6.0. To compare the frequency of psychiatric problems between both samples, we matched the mothers based on their current age using the R ‘MatchIt’ package, with a proportion 1 (genetic sample) to 10 (community sample). We then compared the frequencies of each group using chi-squared tests.

## Results

The socio-demographic characteristics of the mothers of children with CNV are described in Table 1. Depression was the most common psychiatric disorder, affecting 54% (N=146), of the sample (Table 2). Out of those, the majority sought treatment (84%, 121/144), and received medication (74%, 108/145), while 1% (13/132) was hospitalized. Of the 146 mothers reporting depression, 88 indicated the onset, and of those 43/88 (49%) reported their depression started after the birth of their child with a CNV. Anxiety was reported by 31% (N=85), of the sample. Similar to depression, the majority of individuals, sought treatment (73%, 61/83), and 55% (46/84) received medication for their anxiety, while 4% (3/84) was hospitalized. Of the 85 mothers who reported anxiety 49 indicated the onset, and of those 55% (27/49) reported their anxiety started after the birth of their child. Other psychiatric disorders reported included panics (29%), phobias (21%), anorexia and bulimia (12%), other problems (i.e., psychotic or OCD related disorders) (13%), and alcohol/drug problems (6%). We found no evidence that the frequency of depression (OR=0.55, 95% CI=0.11 to 2.27, p=0.43) or anxiety (OR=0.89, 95% CI=00.12 to 4.12, p=0.89) were higher in mothers who also carried a CNV themselves, compared to mothers who did not.

**Table 1.** Socio-demographic characteristics of mothers of children with CNVs

| **Family ethnic background** |  |
| --- | --- |
| European | 81% |
| Mixed | 8% |
| Unknown | 11% |
| **Highest maternal educational qualification** |  |
| Low | 7% |
| Middle | 43% |
| High | 31% |
| Unknown | 19% |
| **Family income** |  |
| <£9,999 | 6% |
| £10,000-£19,999 | 18% |
| £20,000-£39,000 | 31% |
| £40,000-£59,000 | 14% |
| £60,000+ | 17% |
| Unknown | 14% |

**Table 2.** Frequency of psychiatric disorders in Mothers of children with CNV (N=272)

| **Disorder** |  | **After birth of child** |
| --- | --- | --- |
|  | **N(%)** | **N(%)** |
| **Depression** | 146(54%) | 43(49%) |
| Sought treatment | 121(84%) |  |
| Received medication | 108(74%) |  |
| Hospitalized | 13(1%) |  |
| **Anxiety** | 85(31%) | 27(55%) |
| Sought treatment | 61(73%) |  |
| Received medication | 46(55%) |  |
| Hospitalized | 3(4%) |  |
| **Panics** | 77(29%) | 19(49%) |
| Sought treatment | 39(48%) |  |
| Received medication | 31(39%) |  |
| Hospitalized | 2(3%) |  |
| **Phobias** | 56(21%) | 5(16%) |
| Sought treatment | 21(34%) |  |
| Received medication | 12(19%) |  |
| Hospitalized | 0 |  |
| **Anorexia/Bulimia** | 31(12%) | 5 (28%) |
| Sought treatment | 15(48%) |  |
| Received medication | 3(10%) |  |
| Hospitalized | 2(7%) |  |
| **Other problems** | 34(13%) | 7(37%) |
| Sought treatment | 20(53%) |  |
| Received medication | 7(18%) |  |
| Hospitalized | 2(5%) |  |
| **Drink/Drug problems** | 15(6%) | 1(10%) |
| Sought treatment | 3(20%) |  |
| Received medication | 1(7%) |  |
| Hospitalized | 2(13%) |  |
| **Notes:** N varies between instances, as not all participants responded to all items. The percentage of the 'after the birth of child' variable reflects the percentage out of those with the disorder. |  |  |

### Comparisons between mothers of children with CNV and community mothers

We did not have data on age for 5 mothers of children with CNV, therefore, our comparison of the frequency of depression was based on 268 mothers of children with CNV and 2,680 age-matched community mothers. Because the question in the community sample did not correspond exactly to the question asked in the mothers of children with CNV sample (see Methods), we compared the frequency of depression in this group, with all three levels of the depression-related question in the mothers of children with CNV sample (Table 3). Mothers of children with CNV had higher frequency of all the three levels of depression compared to the Mothers-Community sample (p<0.001). Furthermore, mothers of children with CNV reported higher frequency of anorexia, bulimia, alcohol abuse and drug addition, compared to mothers from the community sample.

**Table 3.** Comparisons of psychiatric disorders’ frequencies between mothers of children with CNV and mothers from the community sample, as well as before and after the birth of the child.

|  | **Mothers of children with CNV (N=268)** | **Mothers from the community sample (N=2680)** |  |  |
| --- | --- | --- | --- | --- |
| | **Nsample/Ntotal** | **Nsample/Ntotal** | $\chi^2$ | **p** |
| **Ever Depression** | 142/268 (53%) | 319/2680(13%) | 311.69 | <0.001 |
| Sought treatment | 120/142(45%) |  | 207.74 | <0.001 |
| Medication | 105/142 (39%) |  | 147.2 | <0.001 |
|  | **Mothers of children with CNV (N=246)** | **Mothers from the community sample (N=2460)** |  |  |
| **Anorexia** | 21/245 (9%) | 59/2460 (2%) | 29.4 | <0.001 |
| **Bulimia** | 17/245 (7%) | 61/2460 (3%) | 15.7 | <0.001 |
|  | **Mothers of children with CNV (N=242)** | **Mothers from the community sample (N=2420)** |  |  |
| **Alcohol abuse** | 10/242 (4%) | 21/2420 (1%) | 20.4 | <0.001 |
|  | **Mothers of children with CNV (N=244)** | **Mothers from the community sample (N=2440)** |  |  |
| **Drug addiction** | 7/244 (3%) | 11/2440 (0%) | 19.5 | <0.001 |
| **After the birth of the child\*\*** |  |  |  |  |
|  | **Mothers of children with CNV (N=88)** | **Mothers from the community sample (N=176)** |  |  |
| | **N(Nsample/Ntotal)** | **N(Nsample/Ntotal)** | $\chi^2$ | **p** |
| **Depression** | 43/88(48%) | 77/176(44%) | 0.62 | 0.43 |
|  | **Mothers of children with CNV (N=84)** | **Mothers from the community sample (N=840)** |  |  |
| **Anxiety** | 27/57(55%) | 184/656(30%) | 4.5 | 0.03 |
| Notes: Individuals were matched for their age at the time their child was born. 5 Mothers of children with CNV included in previous tables, did not have data on age, therefore they were not included in this analysis. There was no question available in ALSPAC that we could directly map to the ECHO study related to 'ever had anxiety disorders'. **In the 'After the birth of the child' table, we selected mothers who had a psychiatric diagnosis and also data on whether the diagnosis was before or after the birth of their child. Out of 268 mothers of children with CNV, 88 mothers had complete information on whether their psychiatric diagnosis was before or after the birth of their child. They were matched in a 1:2 ratio with mothers from the community sample, because the total of the mothers from the community sample with data on depression and whether it was before or after the birth of their child was N=863. |  |  |  |  |

The frequencies of depression arising post the birth of the index child were similar between mothers of children with CNV (48%) and mothers from the community sample (44%, p=0.43). There was however evidence of difference in the frequency of anxiety disorders (55% mothers of children with CNV vs. 30% mothers from the community sample, p=0.03).

## Discussion

Our study is the first to compare the frequency of psychiatric problems between mothers of children with CNVs and an aged-matched sample of mothers from a community study. The findings supported our hypothesis. Psychiatric problems including depression, eating disorders, drug abuse, were more frequently reported by the mothers of children with CNV, compared to mothers from the community sample. Studies in the general population (e.g., (Goodman & Gotlib, 1999), as well as in 22q11.2 Deletion Syndrome (Sandini, Schneider, Eliez, & Armando, 2020), have indicated associations between parental and child psychopathology. However, the nature and direction of these associations is not yet clear. For example, it is possible that maternal psychiatric problems may increase the risk of psychiatric problems in the child via shared genetic background or via the family environment. Studies on 22q11.2 Deletion syndrome (22q11.2DS), a pathogenic CNV known for its 20 -old increase risk for schizophrenia, have indicated that environmental factors are related to the child’s difficulties (e.g., (Allen et al., 2014; Shashi et al., 2010)). For example, a study by (Allen et al., 2014) found associations between better parental organisation and fewer child difficulties in children with 22q11.2DS. Maternal psychiatric problems may contribute to a less structured family environment that could in turn contribute to increasing risk of psychiatric problems in the child. On the other hand, it is also possible that the increased physical and mental health problems associated with the CNV in the child, may increase the risk of depression and anxiety in the parent due to the high demands of parental caregiving. Longitudinal studies in families of children with developmental disabilities have indicated that parenting stress and child behaviour problems can also have a bi-directional association (Woodman, Mawdsley, & Hauser-Cram, 2015). Futures studies are needed to examine the relationship between parental psychopathology, family environment and child psychopathology in families of children with CNVs, cross-sectionally as well as longitudinally.

We did not find differences in the frequency of depression and anxiety between mothers who were CNV carriers themselves compared to mothers who were not. This result is likely due to low power given the low sample sizes (38 mothers with a CNV vs. 107 mothers without a CNV). Future studies are needed to better understand how maternal psychopathology may be related to child psychopathology in the context of pathogenic CNVs.

Anxiety disorders were more frequently reported by mothers of children with CNV after the birth of their child with a CNV, compared to mothers from the community sample. These findings are in line with studies providing evidence that bringing up a child with a neurodevelopmental disorder can be taxing on the parent in terms of their emotional (e.g., (Y. Mori et al., 2019)) and physical well-being (e.g., (Laurvick et al., 2006)). Research has identified that mothers with a child with a developmental disability were more likely to experience difficulties with coping with stress (Minnes et al., 2015), reporting worse emotional well-being than women in the general population (Yuka Mori et al., 2018; Totsika et al., 2011). Although we matched the mothers for age, it was not possible to match for Socio Economic Status because of different indices used between the studies. Also, ascertainment differences across the studies as well as the different measures used for the assessment of psychiatric problems in the two groups, may have influenced our results.

In conclusion, we found higher frequencies of depression and a number of other psychiatric problems in the mothers of children with CNVs compared to age-matched mothers from the community sample. We also found that the frequency of anxiety disorder was higher in mothers of children with CNV after the birth of their child with a CNV, compared to mothers from the community sample. Our findings highlight the need for management of the mental health problems, not only of the children with the CNVs, but also their parents.

## Data Availability

Data available upon request.

## Acknowledgements

The authors are extremely grateful to all the families who took part in this study, the midwives for their help in recruiting them, and the whole ALSPAC, ECHO and IMAGINE-ID study teams, which includes interviewers, computer and laboratory technicians, clerical workers, research scientists, volunteers, managers, receptionists, and nurses.

## Data availability

Due to ethical restrictions, data is not available to be shared.

## Notes

**Conflict of interest:** JH, MJO and MVB are funded by a research grant from Takeda Pharmaceuticals outside the scope of the current work.

**Funding:** This research was funded by MRC grant Intellectual Disability and Mental Health: Assessing Genomic Impact on Neurodevelopment (IMAGINE) (MR/N022572/1 and MR/L011166/1; JH, MvdB and MO), a Wellcome Trust Strategic Award ‘Defining Endophenotypes From Integrated Neurosciences (DEFINE) (503147 MO and JH), Medical Research Council Programme Grant (G0800509; MO), the National Institute of Mental Health (NIMH 5UO1MH101724; MvdB and MO), a Wellcome Trust Institutional Strategic Support Fund (ISSF) award (MvdB), the Waterloo Foundation (918-1234; MvdB), the Baily Thomas Charitable Fund (2315/1; MvdB and Health & Care Research Wales (Welsh Government, 507556).

### Competing Interest Statement

JH, MJO, and MVB are funded by a research grant from Takeda Pharmaceuticals outside the scope of the current work.

### Funding Statement

This research was funded by MRC grant Intellectual Disability and Mental Health: Assessing Genomic Impact on Neurodevelopment (IMAGINE) (MR/N022572/1 and MR/L011166/1, JH, MvdB and MO), a Wellcome Trust Strategic Award Defining Endophenotypes From Integrated Neurosciences (DEFINE) (503147 MO and JH), Medical Research Council Programme Grant (G0800509; MO),the National Institute of Mental Health (NIMH 5UO1MH101724; MvdB and MO), a Wellcome Trust Institutional Strategic Support Fund (ISSF) award (MvdB), the Waterloo Foundation (918-1234; MvdB), the Baily Thomas Charitable Fund (2315/1; MvdB and Health & Care Research Wales (Welsh Government, 507556).

### Author Declarations

The ECHO and IMAGINE-ID studies were approved by the National Health Service Wales Research and NHS London Queen Square research ethics committees. Parents provided informed written consent for their participation. Regarding the ALSPAC study, ethical approval was provided by the Local Research Ethics Committees and the ALSPAC's Law and Ethics Committee. Parents provided informed written consent and were allowed to withdraw at any time.

### Summary of Updates

Corrected the p-value of the frequency of anxiety between mothers of children with CNVs compared to mothers from the community sample, and updated abstract.

