## Supplement for "Psychopathology in mothers of children with pathogenic Copy Number Variants"

**IMAGINE-ID Consortium Membership**

^1^ School of Clinical Medicine, University of Cambridge, Cambridge Biomedical Campus, Cambridge, UK

^2^ Medical Research Council Centre for Neuropsychiatric Genetics and Genomics, Division of Psychological Medicine and Clinical Neurosciences, and Neuroscience and Mental Health Research Institute, Cardiff University, Cardiff, UK.

^3^ NIHR BRC Great Ormond Street Institute of Child Health, University College London, London, UK

^4^ Unique – The Rare Chromosome Disorder Support Group, London, UK.

| **Title (if applicable)** | **First Name** | **Surname** | **Institution** |
| --- | --- | --- | --- |
| Dr | Kate | Baker | 1 |
|  | Eleanor | Dewhurst | 1 |
|  | Amy | Lafont | 1 |
| Professor | F Lucy | Raymond | 1 |
|  | Terry | Shirley | 1 |
|  | Hayley | Tilley | 1 |
|  | Husne | Timur | 1 |
|  | Catherine | Titterton | 1 |
|  | Neil | Walker | 1 |
|  | Sarah | Wallwork | 1 |
|  | Francesca | Wicks | 1 |
| Dr | Zheng | Ye | 1 |
|  | Marie | Erwood | 1 |
|  | Sophie | Andrews | 2 |
|  | Philippa | Birch | 2 |
|  | Samantha | Bowen | 2 |
|  | Karen | Bradley | 2 |
|  | Aimee | Challenger | 2 |
| Dr | Samuel | Chawner | 2 |
| Dr | Andrew | Cuthbert | 2 |
| Professor | Jeremy | Hall | 2 |
| Professor | Peter | Holmans | 2 |
|  | Sarah | Law | 2 |
|  | Nicola | Lewis | 2 |
|  | Sinead | Morrison | 2 |
|  | Hayley | Moss | 2 |
| Professor Sir | Michael | Owen | 2 |
|  | Sinead | Ray | 2 |
|  | Matthew | Sopp | 2 |
|  | Molly | Tong | 2 |
| Professor | Marianne | van den Bree | 2 |
|  | Nadia | Coscini | 3 |
|  | Sarah | Davies | 3 |
|  | Spiros | Denaxas | 3 |
|  | Hayley | Denyer | 3 |
|  | Nasrtullah | Fatih | 3 |
|  | Manoj | Juj | 3 |
|  | Ellie | Kerry | 3 |
|  | Anna | Lucock | 3 |
| Dr | William | Mandy | 3 |
|  | Frida | Printzlau | 3 |
| Professor | David | Skuse | 3 |
| Dr | Ramya | Srinivasan | 3 |
| Dr | Susan | Walker | 3 |
|  | Alice | Watkins | 3 |
| Dr | Jeanne | Wolstencroft | 3 |
| Dr | Beverly | Searle | 4 |
| Dr | Anna | Pelling | 4 |

IMAGINE-ID Clinical Collaborators

| **Title (if applicable)** | **First Name** | **Surname** | **Shorthand Institution** | **Genetics service** | **Hospital Trust** |
| --- | --- | --- | --- | --- | --- |
| Dr | John | Dean | Aberdeen | Aberdeen Royal Infirmary Genetics Service | NHS GRAMPIAN |
| Dr | Lisa | Robertson | Aberdeen | Aberdeen Royal Infirmary Genetics Service | NHS GRAMPIAN |
| Dr | Denise | Williams | Birmingham | West Midlands Regional Genetics Service | BIRMINGHAM WOMEN'S NHS FOUNDATION TRUST |
| Dr | Alan | Donaldson | Bristol | Bristol Clinical Genetics Service | UNIVERSITY HOSPITALS BRISTOL NHS FOUNDATION TRUST |
| Professor | Lucy | Raymond | Cambridge | East Anglian Medical Genetics Service | CAMBRIDGE UNIVERSITY HOSPITALS NHS FOUNDATION TRUST |
| Dr | Annie | Procter | Cardiff | All Wales Regional Genetics Service | CARDIFF AND VALE UNIVERSITY LHB |
| Dr | Jonathan | Berg | Dundee | Ninewells Hospital Dundee Genetics Service | NHS TAYSIDE |
|  | Yanick | Crow | Edinburgh | Western General Hospital Edinburgh Genetics Service | NHS LOTHIAN |
| Professor | Anne | Lampe | Edinburgh | Western General Hospital Edinburgh Genetics Service | NHS LOTHIAN |
| Dr | Julia | Rankin | Exeter | Peninsula Genetics Service | ROYAL DEVON AND EXETER NHS FOUNDATION TRUST |
| Dr | Shelagh | Joss | Glasgow | Glasgow Genetics Centre | NHS GREATER GLASGOW & CLYDE |
| Professor | Lyn | Chitty | GOSH | London North East Thames Regional Genetics Service - Clinical Genetics | GREAT ORMOND STREET HOSPITAL FOR CHILDREN NHS FOUNDATION TRUST |
| Professor | Frances | Flinter | Guy's | London Guy's Hospital Genetic Centre | GUY'S AND ST THOMAS' NHS FOUNDATION TRUST |
| Dr | Muriel | Holder | Guy's | London Guy's Hospital Genetic Centre | GUY'S AND ST THOMAS' NHS FOUNDATION TRUST |
| Dr | Alison | Kraus | Leeds | Yorkshire Regional Genetics Service - Clinical Genetics | LEEDS TEACHING HOSPITALS NHS TRUST |
| Dr | Julian | Barwell | Leicester | Leicestershire Genetics Centre | UNIVERSITY HOSPITALS OF LEICESTER NHS TRUST |
| Dr | Pradeep | Vasudevan | Leicester | Leicestershire Genetics Centre | UNIVERSITY HOSPITALS OF LEICESTER NHS TRUST |
| Dr | Astrid | Weber | Liverpool | Cheshire & Merseyside Regional Genetic Service | LIVERPOOL WOMEN'S NHS FOUNDATION TRUST |
| Dr | William | Newman | Manchester | Manchester Centre for Genomic Medicine | CENTRAL MANCHESTER UNIVERSITY HOSPITALS NHS FOUNDATION TRUST |
| Dr | Miranda | Splitt | Newcastle | Northern Genetics Service | THE NEWCASTLE UPON TYNE HOSPITALS NHS FOUNDATION TRUST |
| Dr | Virginia | Clowes | North West Thames | London North West Thames Regional Genetics Service | NORTH WEST LONDON HOSPITALS NHS TRUST |
| Dr | Fleur | van Dijk | North West Thames | London North West Thames Regional Genetics Service | NORTH WEST LONDON HOSPITALS NHS TRUST |
| Dr | Rachel | Harrison | Nottingham | Nottingham Regional Genetics Service | NOTTINGHAM UNIVERSITY HOSPITALS NHS TRUST |
| Dr | Usha | Kini | Oxford | Oxford Genetics Service | OXFORD UNIVERSITY HOSPITALS NHS TRUST |
| Dr | Oliver | Quarrell | Sheffield | Sheffield Genetic Services | SHEFFIELD CHILDREN'S NHS FOUNDATION TRUST |
| Dr | Diana | Baralle | Southampton | Wessex Clinical Genetics Service | UNIVERSITY HOSPITAL SOUTHAMPTON NHS FOUNDATION TRUST |
| Dr | Sahar | Mansour | St George's | London South West Thames Regional Genetics Service | ST GEORGE'S HEALTHCARE NHS FOUNDATION TRUST |
